# Diagnostic Accuracy and Risk Stratification of a Trauma Risk Assessment Tool Among those with Fall Injuries

**DOI:** 10.1101/2023.06.19.23291576

**Authors:** Oluwaseun John Adeyemi, Sanjit Konda, Charles DiMaggio, Corita R. Grudzen, Ashley Pfaff, Garrett Esper, Mauricio Arcila-Mesa, Allison M. Cuthel, Helen Poracky, Polina Meyman, Ian Wittman, Joshua Chodosh

**Affiliations:** Ronald O. Perelman Department of Emergency Medicine, NYU Grossman School of Medicine, New York, USA; Department of Orthopedic Surgery, NYU Grossman School of Medicine, New York, USA; Department of Surgery, NYU Grossman School of Medicine, New York, USA; Department of Population Health, NYU Grossman School of Medicine, New York, USA; Department of Medicine, Memorial Sloan Kettering Cancer Center, USA; Department of Medicine, NYU Grossman School of Medicine, New York, USA; Department of Trauma, NYU Langone Hospital, New York, USA; Medicine Service, Veterans Affairs New York Harbor Healthcare System, New York, NY, USA

**Keywords:** Diagnostic Accuracy, Falls, Mortality, Risk-triage, Geriatrics

## Abstract

**Aim:** The Score for Trauma Triage in the Geriatric and Middle-Aged (STTGMA) is an injury risk-triage tool. This study aims to validate the STTGMA’s accuracy in predicting fall-related mortality among geriatric trauma patients.

**Methods:** Using a retrospective cohort design, we selected 5,791 geriatric trauma patients (aged 55 years and older) from a single institutional trauma database (2017-2021). The outcome variable was fatal fall injury, measured as a binary variable. The predictor variable was the STTGMA score, measured as a continuous variable and a four-level categorical variable. We report the predictive accuracy (95% confidence interval (CI)) of the STTGMA. We further assessed the relationship between the STTGMA risk categories and hospital length of stay and time-to-death by performing multivariable quantile regression and time-varying Cox proportional hazard analyses, respectively.

**Results:** A total of 122 patients (2.1%) died during admission and the median hospital length of stay was 2 days. STTGMA exhibited 84% (95% CI: 75.6 – 92.0) accuracy in predicting in-hospital fall-related mortality. Compared to the minimal risk category, geriatric trauma patients classified as low, moderate, and high risks each had significantly longer hospital stays and adjusted mortality risks, in a dose-response pattern.

**Conclusion:** STTGMA can accurately predict in-hospital mortality and risk-stratify the length of stay and the time to death among geriatric patients with fall injuries.

## Introduction

Falls are the leading cause of injury-related mortality among US older adults (aged 65 years and older) and every day, approximately 90 older adults die from fall-related injuries.(1) Between 2007 and 2016, the age-adjusted fatal fall rate among older adults increased by approximately 30 percent, from 47 to 62 deaths per 100,000 population, with rates highest among adults 85 years and older.(2, 3) With 10,000 US adults turning 65 years daily,(4) the volume of fall-related injuries is expected to increase. Currently, fall-related injuries account for approximately 800,000 emergency department (ED) visits annually, and the US spends over $750 million yearly on fatal fall injuries.(3)

Despite fall-related injuries accounting for two-thirds of geriatric trauma injuries,(5) fall-related injuries are more likely to be under-triaged.(6) The underestimation of fall-related injury severity among older adults further predisposes these patients to increased morbidity and mortality.(6-8) The commonly used injury triage tools such as the Revised Trauma Scale,(9) the Injury Severity Scale and the Trauma and Injury Severity Score have exhibited poor diagnostic accuracy among older adults.(10-12) Trauma-specific scales such as the Ohio geriatric guideline have demonstrated 52 to 65% accuracy in predicting level I and II trauma center transfer while the Manchester triage guideline showed a 74% accuracy in predicting geriatric trauma mortality.(13, 14). However, these trauma scales have not been used specifically for fall-related mortality.

The Score for Trauma Triage in Geriatric and Middle Age (STTGMA) is a risk triage tool that predicts mortality risk using age, injury severity characteristics, and background comorbidities.(15) The STTGMA risk triage tool has primarily been used in the geriatric orthopedic trauma population to predict mortality from fracture-related injuries.(16, 17) It has also been implemented to predict in-hospital disposition, the need for a blood transfusion, and the cost of care among orthopedic trauma patients.(18-22) Across these studies, the STTGMA risk triage tool has demonstrated 74 to 94% accuracy in predicting in-hospital mortality.(16, 17)

Assessing STTGMA’s predictive accuracy for fall-related mortality may validate the usability of the instrument for the care of geriatric trauma patients. No study has assessed the sensitivity, specificity, and diagnostic accuracy of the STTGMA risk-triage tool among geriatric trauma patients with fall injuries. Additionally, investigating the relationship between STTGMA and other clinical measures of fall-related mortality such as hospital length of stay and time-to-death may further enhance its in-hospital utility. This study aims, therefore, to assess the diagnostic accuracy of the STTGMA risk triage tool in predicting in-hospital mortality and to assess the association between the STTGMA-derived risk categories and hospital length of stay and time-to-death from fall-related injuries.

## Methods

### Study Design and Setting

Using a retrospective cohort design, we pooled trauma registry data between 2017 and 2021 from a single Level-I trauma center that serves a racially diverse community in New York. Data were extracted in June 2022 and the data were analyzed between January and March 2023. This study is among the validation studies focused on exploring the diagnostic accuracies of a novel scoring tool for geriatric trauma patients. We obtained Institutional Review Board (IRB) approval from the New York University Langone Health IRB (i20_01316_MOD05). The authors did not have access to information that could identify individual participants during or after data collection. The results we present follow the reporting guideline of the Standards for Reporting Diagnostic Accuracy.(23)

### Inclusion and Exclusion Criteria

We selected adults, 55 years and older, who presented to the ED with traumatic injuries (N=7,634) (Figure 1). We included patients between the ages of 55 and 64 because trauma-related mortality significantly increases after age 55.(24) We excluded patients whose injuries were not related to falls (n=1,102). Duplicate entries (recurrent within-patient ED visits) were identified and we used the most recent encounter (n=740). We performed a listwise deletion for variables with missing entries when the proportion was less than 1% (variable: sex; n=1 (0.0%)). The final analysis included 5,791 adults aged 55 years and older who presented to the ED with fall-related injuries.

**Figure 1:**
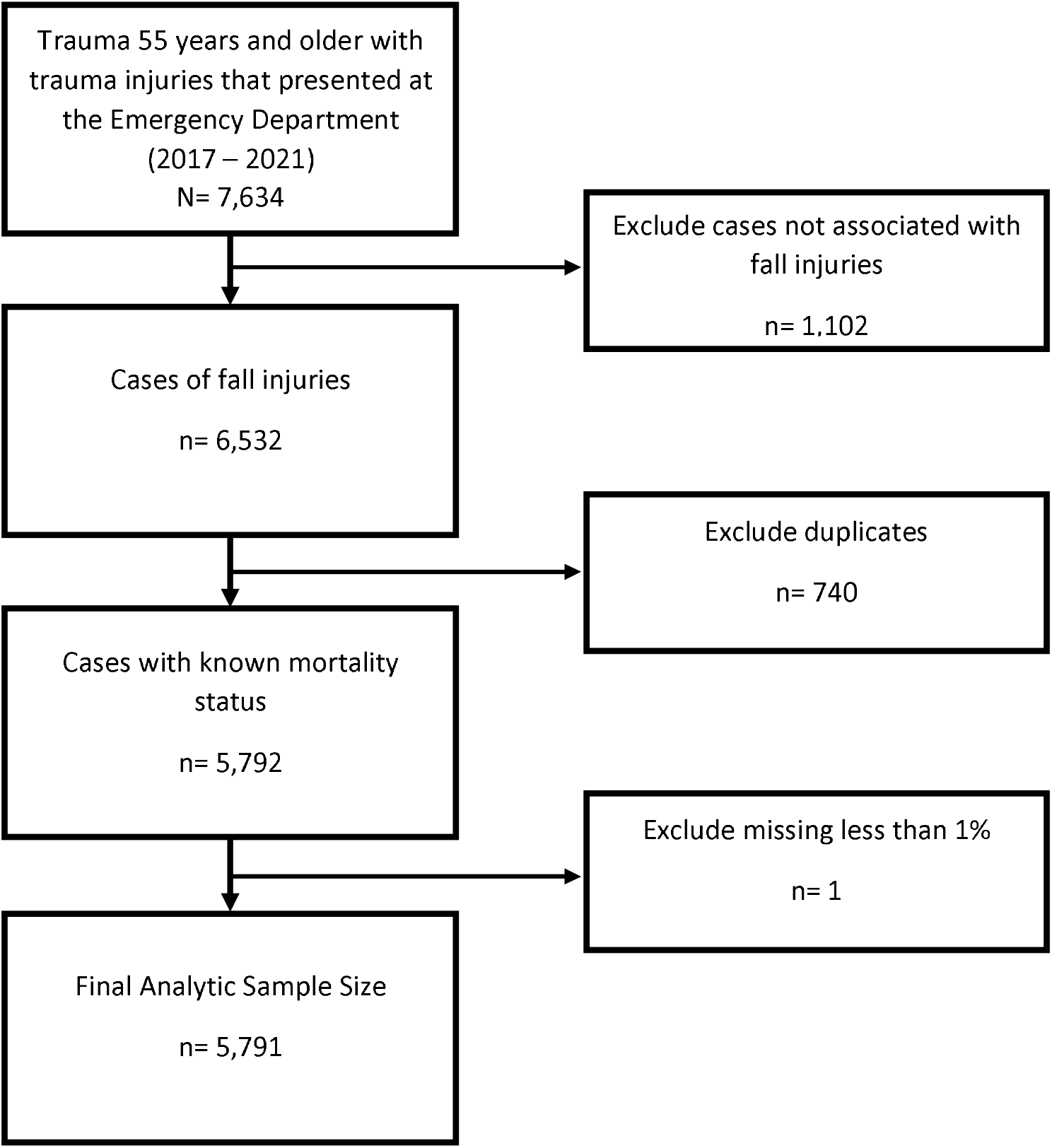
Data selection steps

### Handling of Missing Data

Three variables had missing values less than 10% - race/ethnicity (n=96, 1.7%), body mass index (n=295 (5.1%)), and Glasgow Coma Scale score (n=539, (9.3%)). We performed multiple imputations after confirming that the missing pattern was missing at random using Little’s test. (25-27) The multiple imputations used the multivariate normal regression with 100 iterations and the imputed value represents the average across the 100 iterations.(28, 29)

### Data

From the analytic dataset (n=5,791), we created two separate pools of data: (1) the training dataset (n=4,054) and (2) the test dataset (n=1,737). The training dataset was used for the Receiver Operating Curve (ROC) analysis while the test dataset was used for the internal validation analysis. The split was done in a 70:30 ratio, using simple random sampling with no replacement.

### Outcome Variables

The primary outcome measure of interest was in-hospital death. We defined in-hospital death as death occurring either in the ED or during the index admission. In-hospital death was defined as a binary variable (yes or no) and this variable served as the outcome of interest for the ROC and internal validation analyses.

The secondary outcome measures of interest were hospital length of stay and the time to death from fall-related injuries. The hospital length of stay was defined as the duration from hospital admission to final hospital disposition. The final hospital disposition was defined as discharge to home or facility, transfer to another hospital or facility, or death. Hospital length of stay was measured as a continuous variable. Time to death from fall-related injury was defined as the time from admission following the injury to the time of in-hospital death. Patients who were alive at the time of discharge were considered as having right censored events.

### Predictor Variable: STTGMA

The predictor variables were the STTGMA risk-triage score and the STTGMA risk categories. The STTGMA risk-triage score was computed using an online Excel calculator.(15) First, we determined if the injury was high or low-energy mechanisms. High-energy falls were falling from heights greater than two meters or collisions with an automobile while low-energy falls were falling from a standing height or lower. Thereafter, we calculated the STTGMA score using the patient’s age, Glasgow Coma Scale score, Charlson Comorbidity Index, and the Abbreviated Injury Severity scores of the head and neck, chest, and extremity and pelvis. Age and Charlson Comorbidity Index were measured as continuous variables. Glasgow Coma Scale score, typically categorized as mild (13 - 15), moderate (8 - 12), and severe (3 to 7) traumatic brain injury was measured as a continuous variable ranging from 3 to 15. Abbreviated Injury Severity, typically categorized as no injury, minor, moderate, serious, severe, critical, or maximal injuries, was scored from 0 to 6, respectively. The STTGMA score is computed as a logarithmic function of these four measures and has a score ranging from 0 to 1. We report the score in percentages, with higher values representing increased injury severity. We generated STTGMA risk categories – minimal (0 – 50%), low (51 – 80%), moderate (81 – 95%), and high (greater than 95%), using the percentile distribution of scores.

### Control Variables

We did not control for any additional variables in the ROC and internal validation analyses. For the secondary analysis, we controlled for sex, race/ethnicity, and body mass index. Sex was defined as male or female. Race/ethnicity was defined as non-Hispanic White, non-Hispanic Black, Hispanic, Asian, and other races. Body mass index was defined as normal weight (18.0 – 24.9kg/m_2_), underweight (<18.0 kg/m_2_), overweight (25.0 – 29.9kg/m_2_), and obese (≥30.0kg/m_2_).

### Data Analysis

We reported the frequency distribution and summary statistics in the complete, training, and test datasets and reported the differences across the training and test datasets using the chi-square test, independent sample t-test, and Mann-Whitney U test as appropriate. We computed the median differences in the hospital length of stay and assessed the difference across the predictor and control variables using the Mann-Whitney U test and the Kruskal Wallis test as appropriate. Also, we computed the case fatality rates across the predictor and control variables and assessed differences across the variables using the log-rank test.

For the ROC and internal validation analyses, we first assessed the logarithmic relationship between STTGMA risk-triage scores and in-hospital death and generated the predicted estimates. Thereafter, we created the area under the ROC (AUROC) using the sensitivity on the y-axis and the 1-specificity on the x-axis and reported the accuracy and the 95% confidence interval (CI). AUROC values between 0.7 and 0.8 are interpreted as acceptable, 0.8 to 0.9 as excellent, and higher than 0.9 as outstanding.(30) Also, we computed the Youden index - a measure of a tool’s effectiveness as a diagnostic marker.(23) The Youden index ranges from 0 to 100% and values of 50% or higher are considered adequate.(31)

For the secondary analysis, we performed a quantile regression analysis to assess the association between the STTGMA risk categories and hospital length of stay. We reported the unadjusted and adjusted median differences and the 95% CI. We also performed a time-varying Cox proportional hazard regression analysis to assess the differences between the STTGMA risk categories and the time-to-death from fall-related injuries. We selected a time-varying model since the test of proportionality of strata was significant, evidenced by the crossing of the log-log plot of the STTGMA risk categories, and the significant Schoenfeld residual proportional hazard test.(32) We reported the unadjusted and adjusted hazard risk ratio (mortality risk ratio) and the 95% CI.

## Results

### Descriptive Characteristics

Among the 5,791 adults who sustained fall injuries, the mean (SD) age was 77.5 (11.4) years (Table 1). The population was predominantly female (63%) and non-Hispanic White (63%). Most of the patients were of either normal weight (39%) or overweight (38%). The injury mechanism was low energy (80%) and 95% of the population had mild traumatic brain injuries. Overall, 45% of the population had no associated comorbidities, and the proportion with minor injuries to the head, chest, and extremities were 67%, 94%, and 52%, respectively. The median (Q1, Q3) STTGMA risk score was 1.3% (0.7%, 2.4%). The median (Q1, Q3) hospital length of stay was 2 (0.0, 5.0) days. There was a total of 122 (2.1%) in-hospital deaths. There were no significant differences in the demographic, injury, and hospital disposition characteristics in the training and test datasets. There were significant differences in the hospital length of stay by sex (p = 0.004), race/ethnicity (p < 0.001), body mass index (p < 0.001), and STTGMA risk categories (p<0.001) (Table 2). Also, there were significant differences in the fall case fatality rates by sex and STTGMA risk categories.

**Table 1:**
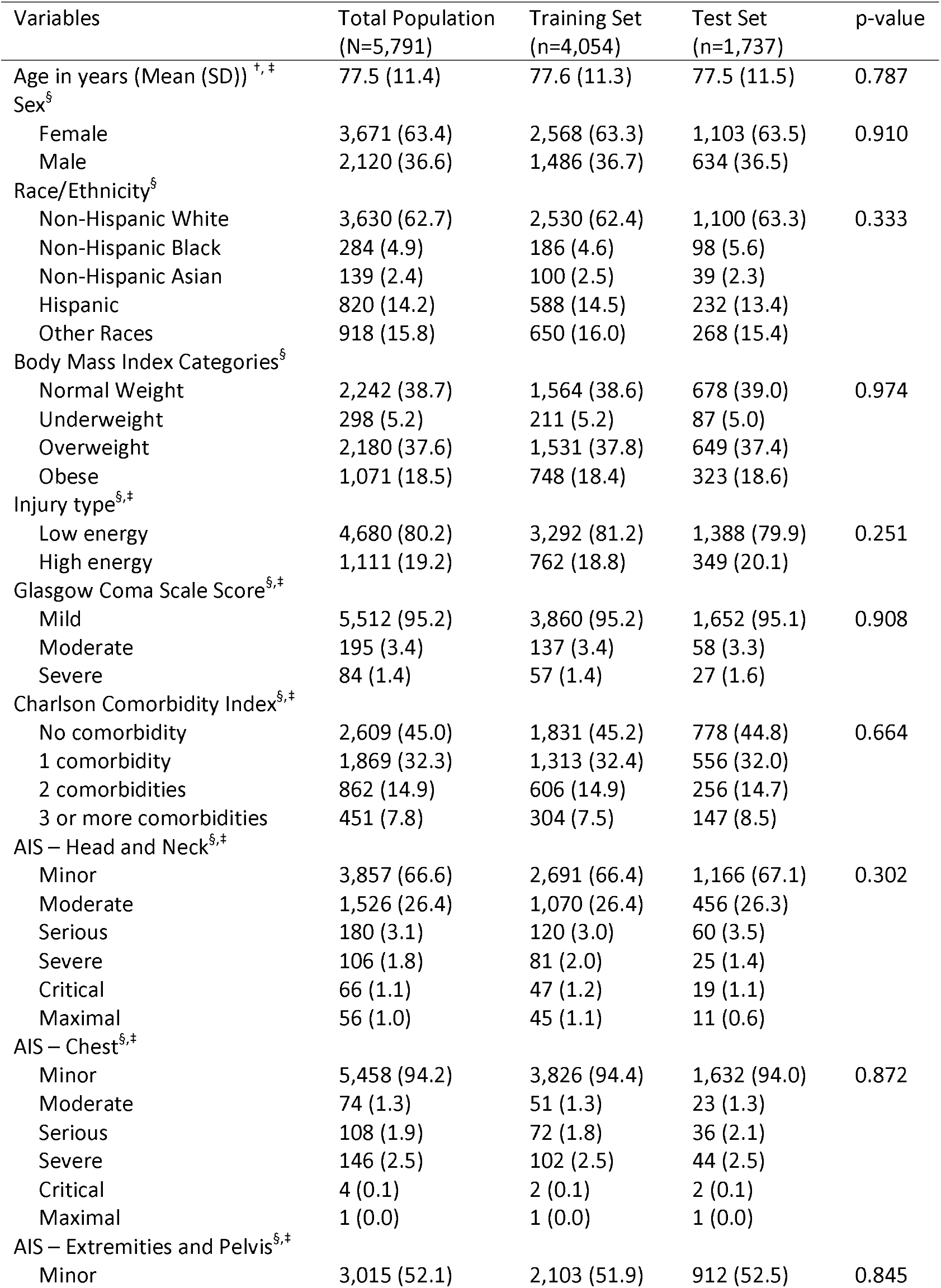
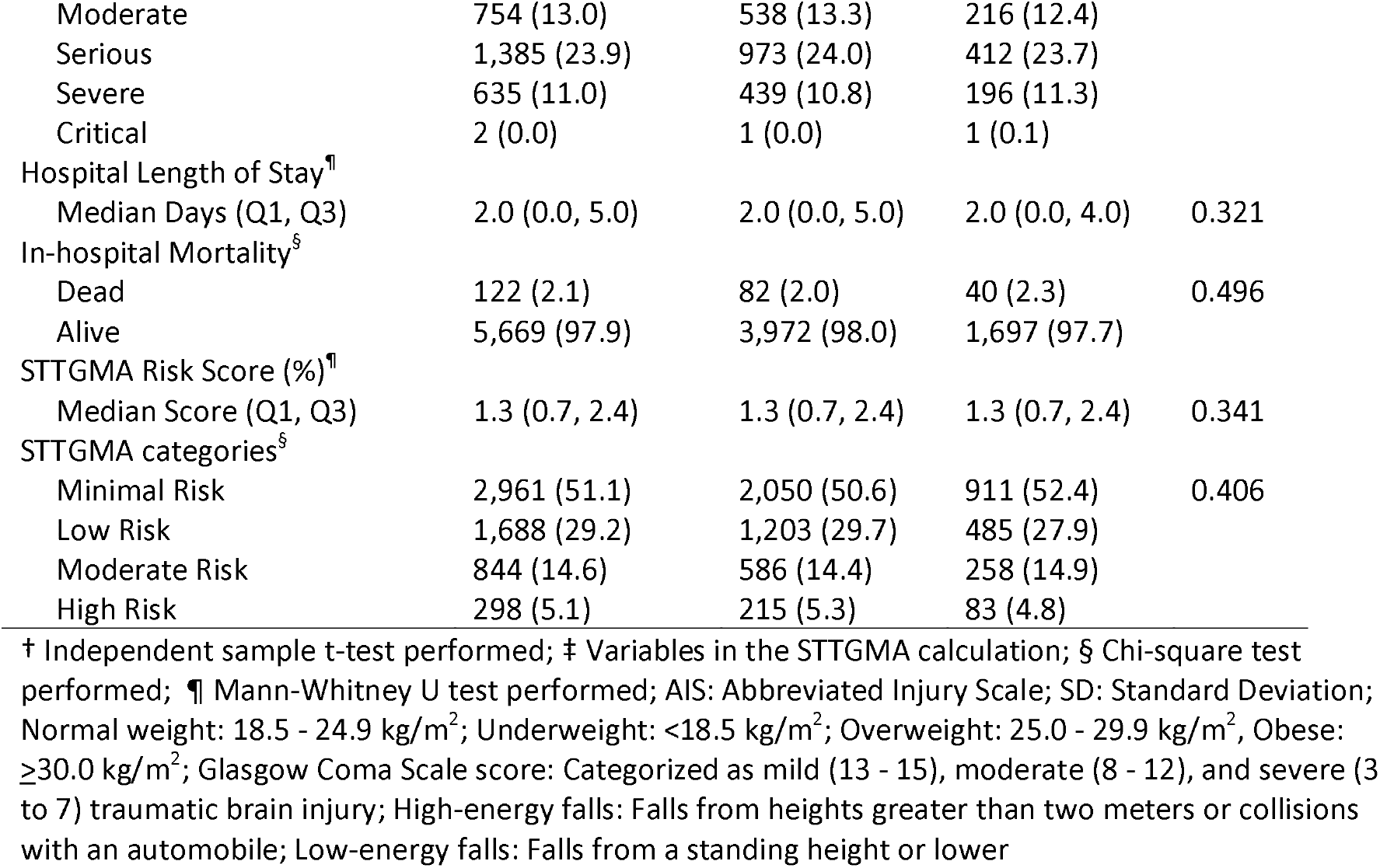
Demographic and injury characteristics of the study population in the training and test datasets among middle age and geriatric trauma patients involved in fall injuries (N=5,791)

**Table 2:**
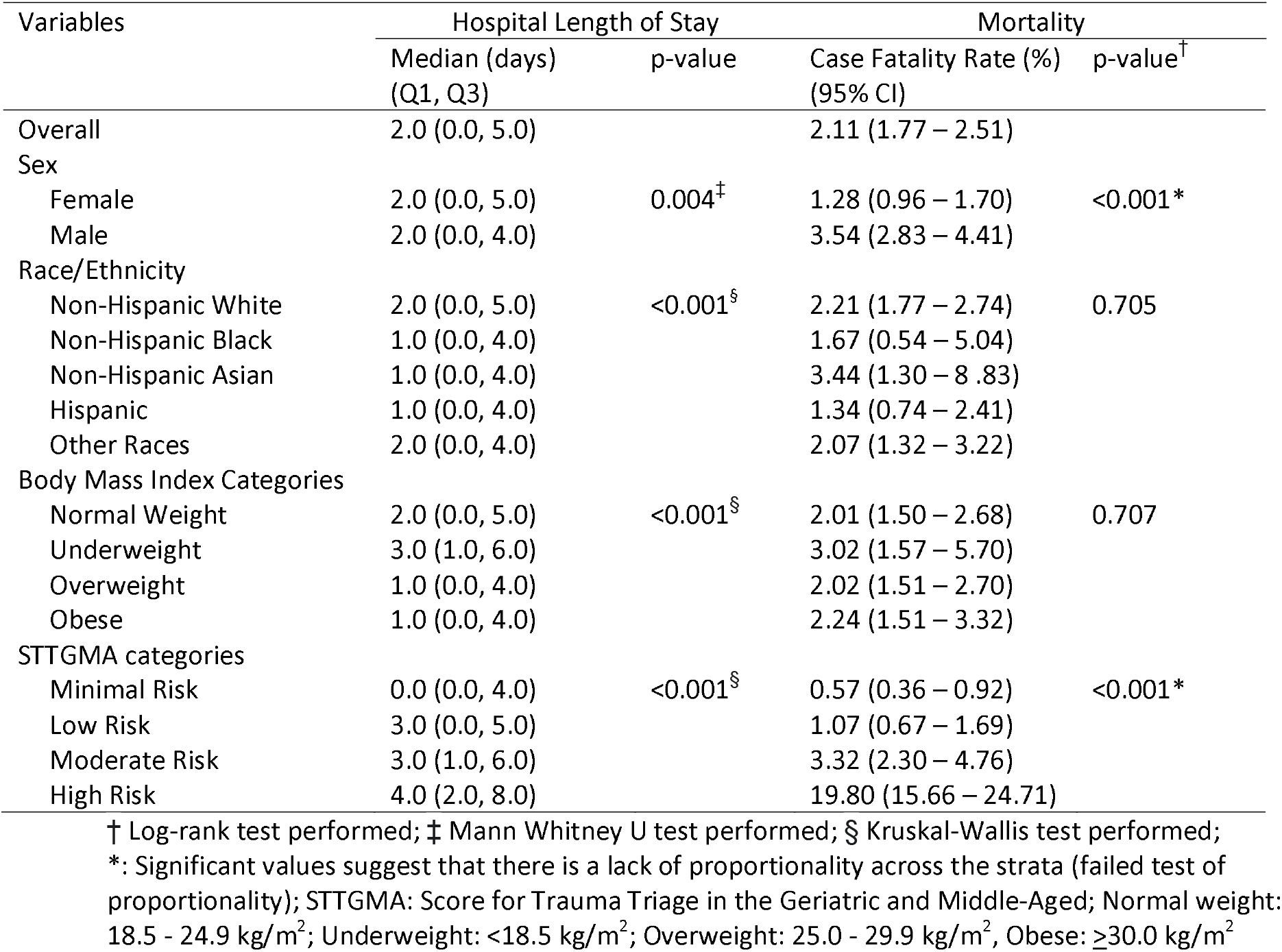
Summary statistics of the hospital length of stay and incidence rates of fatal fall injuries across the study population (N=5,791)

| Variables | Hospital Length of Stay |  | Mortality |  |
| --- | --- | --- | --- | --- |
|  | Median (days)<br>(Q1, Q3) | p-value | Case Fatality Rate (%)<br>(95% CI) | p-value<sup>†</sup> |
| Overall | 2.0 (0.0, 5.0) |  | 2.11 (1.77 – 2.51) |  |
| Sex |  |  |  |  |
| Female | 2.0 (0.0, 5.0) | 0.004<sup>‡</sup> | 1.28 (0.96 – 1.70) | <0.001* |
| Male | 2.0 (0.0, 4.0) |  | 3.54 (2.83 – 4.41) |  |
| Race/Ethnicity |  |  |  |  |
| Non-Hispanic White | 2.0 (0.0, 5.0) | <0.001<sup>§</sup> | 2.21 (1.77 – 2.74) | 0.705 |
| Non-Hispanic Black | 1.0 (0.0, 4.0) |  | 1.67 (0.54 – 5.04) |  |
| Non-Hispanic Asian | 1.0 (0.0, 4.0) |  | 3.44 (1.30 – 8.83) |  |
| Hispanic | 1.0 (0.0, 4.0) |  | 1.34 (0.74 – 2.41) |  |
| Other Races | 2.0 (0.0, 4.0) |  | 2.07 (1.32 – 3.22) |  |
| Body Mass Index Categories |  |  |  |  |
| Normal Weight | 2.0 (0.0, 5.0) | <0.001<sup>§</sup> | 2.01 (1.50 – 2.68) | 0.707 |
| Underweight | 3.0 (1.0, 6.0) |  | 3.02 (1.57 – 5.70) |  |
| Overweight | 1.0 (0.0, 4.0) |  | 2.02 (1.51 – 2.70) |  |
| Obese | 1.0 (0.0, 4.0) |  | 2.24 (1.51 – 3.32) |  |
| STTGMA categories |  |  |  |  |
| Minimal Risk | 0.0 (0.0, 4.0) | <0.001<sup>§</sup> | 0.57 (0.36 – 0.92) | <0.001* |
| Low Risk | 3.0 (0.0, 5.0) |  | 1.07 (0.67 – 1.69) |  |
| Moderate Risk | 3.0 (1.0, 6.0) |  | 3.32 (2.30 – 4.76) |  |
| High Risk | 4.0 (2.0, 8.0) |  | 19.80 (15.66 – 24.71) |  |

† Log-rank test performed; ‡ Mann Whitney U test performed; § Kruskal-Wallis test performed; \*: Significant values suggest that there is a lack of proportionality across the strata (failed test of proportionality); STTGMA: Score for Trauma Triage in the Geriatric and Middle-Aged; Normal weight: 18.5 - 24.9 kg/m<sup>2</sup>; Underweight: <18.5 kg/m<sup>2</sup>; Overweight: 25.0 - 29.9 kg/m<sup>2</sup>, Obese: ≥30.0 kg/m<sup>2</sup>

### ROC Analysis and Internal Validation

Using the training dataset, a unit increase in STTGMA risk score was associated with an 8% increased odds of fatal fall injury (OR: 1.08; 95% CI: 1.06 - 1.09). STTGMA risk score demonstrated 81% accuracy in predicting in-hospital mortality (95% CI: 75.7 - 86.5) (Figure 2A). The sensitivity and specificity of the STTGMA risk score were 80% and 51%, respectively, and the Youden index was 52%. The internal validation, using the test dataset, showed that a unit increase in STTGMA risk score was associated with a 16% increased odds of death after a fall injury (OR: 1.16; 95% CI: 1.12 - 1.20). STTGMA risk score demonstrated 84% accuracy in predicting in-hospital mortality (95% CI: 75.6 - 92.0) with internal validation (Figure 2B). The sensitivity and specificity of the STTGMA risk scores were 83% and 52%, respectively, and the Youden index was 60%.

**Figure 2:**
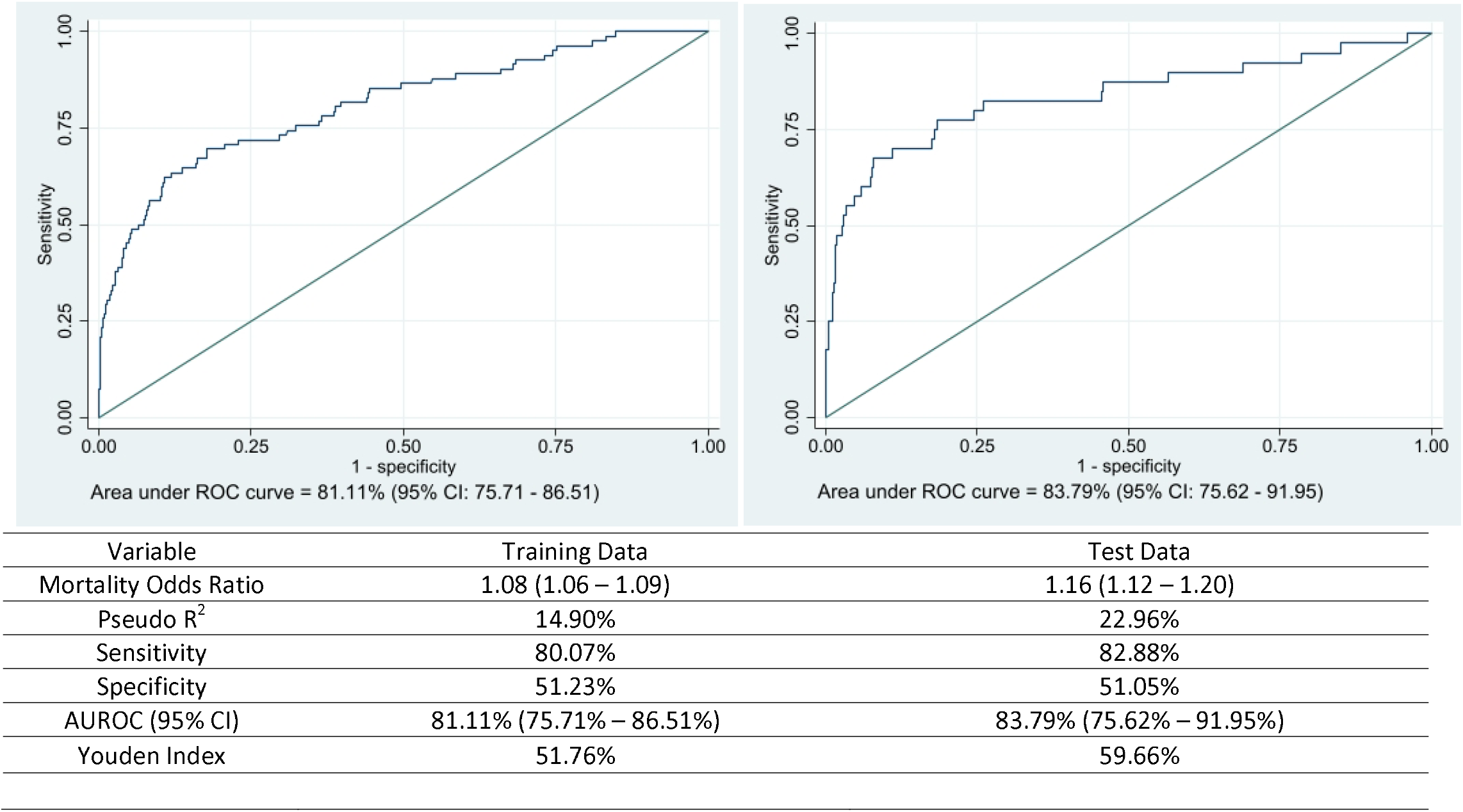
Summary of the diagnostic accuracy of the Score for Trauma Triage in the Geriatric and Middle-Aged (STTGMA) in accessing in-hospital mortality among adults 55 years and older with fall injuries *Youden Index: Ranges from 0 to 100% assuming sensitivity and specificity are of equal value; the higher the value, the better the discriminant property, and values of 50% or higher are acceptable; **AUROC ranging from 0.7 to 0.8 is acceptable, 0.8 to 0.9 is excellent, and values higher than 0.9 as outstanding; AUROC: Area Under Receiver Operating Characteristic

### Hospital Length of Stay

We report the unadjusted median change in the hospital length of stay across the demographic characteristics and STTGMA risk categories (Table 3). In the adjusted models, non-Hispanic Black patients (Adjusted Median Difference (aMD): -1.0; 95% CI: -1.5 – -0.5) and Hispanic patients (aMD: -1.0; 95% CI: -1.3 – -0.7) had shorter hospital length of stays compared to those who were non-Hispanic White. Patients who were underweight had longer lengths of stay compared to those who were of normal weight (aMD: 1.0; 95% CI: 0.5 – 1.5). Compared to the minimal risk category, patients in the low (95% CI: 1.8 – 2.2) and moderate (1.7 – 2.3) risk categories had a two-day median increase in their hospital length of stay while those classified as high risk (95% CI: 2.5 – 3.5) had a three-day median increase in their hospital length of stay.

**Table 3:** Summary of the unadjusted and adjusted quantile regression and time-varying Cox proportional hazard regression assessing the changes in hospital length of stay and time-to-death among the study population (N=5,791)

| Variables | Hospital Length of Stay<sup>†</sup> |  | Mortality Risk<sup>‡</sup> |  |
| --- | --- | --- | --- | --- |
|  | Unadjusted Coefficient (95% CI) | Adjusted Coefficient (95% CI) | Unadjusted Hazard Ratio (95% CI) | Adjusted Hazard Ratio<sup>##</sup> (95% CI) |
| STTGMA categories |  |  |  |  |
| Minimal Risk | Ref | Ref | Ref<sup>§</sup> | Ref<sup>§</sup> |
| Low Risk | **3.00 (2.72 – 3.28)** | **2.00 (1.76 – 2.24)** | **2.17 (1.03 – 4.57)** | **2.36 (1.11 – 5.01)** |
| Moderate Risk | **3.00 (2.64 – 3.35)** | **2.00 (1.69 – 2.31)** | **6.89 (3.18 – 14.92)** | **7.37 (3.38 – 16.09)** |
| High Risk | **4.00 (3.45 – 4.55)** | **3.00 (2.52 – 3.48)** | **32.31 (14.35 – 72.77)** | **33.77 (14.87 – 76.69)** |
| Sex |  |  |  |  |
| Female | Ref | Ref | Ref<sup>§</sup> | Ref<sup>§</sup> |
| Male | 0.00 (-0.25 – 0.25) | 0.00 (-0.21 – 0.21) | **2.21 (1.31 – 3.73)** | **2.06 (1.38 – 3.09)** |
| Race/Ethnicity |  |  |  |  |
| Non-Hispanic White | Ref | Ref | Ref | Ref |
| Non-Hispanic Black | **-1.00 (-1.91 – -0.09)** | **-1.00 (-1.48 – -0.52)** | 0.61 (0.19 – 1.95) | 0.51 (0.12 – 2.10) |
| Non-Hispanic Asian | -1.00 (-2.27 – 0.27) | **-1.00 (-1.67 – -0.33)** | 1.47 (0.54 – 4.03) | 1.19 (0.36 – 3.94) |
| Hispanic | **-1.00 (-1.57 – -0.43)** | **-1.00 (-1.30 – -0.70)** | 0.74 (0.38 – 1.43) | 0.73 (0.35 – 1.53) |
| Other Races | 0.00 (-0.54 – 0.54) | 0.00 (-0.29 – 0.29) | 0.97 (0.58 – 1.62) | 1.04 (0.54 – 2.02) |
| Body Mass Index Categories |  |  |  |  |
| Normal Weight | Ref | Ref | Ref | Ref |
| Underweight | **1.00 (0.44 – 1.56)** | **1.00 (0.52 – 1.48)** | 1.37 (0.66 – 2.82) | 1.61 (0.78 – 3.35) |
| Overweight | **-1.00 (-1.27 – -0.73)** | 0.00 (-0.24 – 0.24) | 1.08 (0.69 – 1.69) | 1.20 (0.77 – 1.89) |
| Obese | **-1.00 (-1.34 – -0.66)** | 0.00 (-0.29 – 0.29) | 1.30 (0.77 – 2.19) | 1.69 (0.97 – 2.95) |

Significant relationships in bold; † Significant result exists if the confidence interval does not cross zero; ‡ Significant result exists if the confidence interval does not cross one.; § Time-varying Cox proportional hazard model performed; ##Interaction of time X male and time X STTGMA risk categories created in the time-varying Cox proportional multivariable regression model; STTGMA: Score for Trauma Triage in the Geriatric and Middle-Aged; Normal weight: 18.5 - 24.9 kg/m<sup>2</sup>; Underweight: <18.5 kg/m<sup>2</sup>; Overweight: 25.0 - 29.9 kg/m<sup>2</sup>, Obese: ≥30.0 kg/m<sup>2</sup>

### Time to Death

We report the unadjusted mortality risk ratio (hazard risk) across demographic characteristics and STTGMA risk categories (Table 3). In the adjusted models, male patients had two times the mortality risk compared to female patients (adjusted Hazard Ratio (aHR): 2.1; 95% CI: 1.4 – 3.1). Patients classified as having low risk had 2.4 times (95% CI: 1.1 – 5.0) the mortality risk compared to those classified as minimal risk and the mortality risk ratio exponentially increased to 7.4 (95% CI: 3.4 – 16.1) and 33.8 (95% CI: 14.9 – 76.7) in the moderate and high-risk categories.

## Discussion

This study provides a validated fall injury triage tool that can accurately predict mortality risk. We report that among adults 55 years and older with fall injuries, the STTGMA demonstrated high accuracy in predicting fatal fall injuries. Using data from a single level I trauma center, the fall case fatality rate and time-to-death exponentially increased in a dose-response pattern across the STTGMA-derived risk categories. Also, STTGMA risk classifies the hospital length of stay into three non-overlapping categories of minimal risks, low to moderate risks, and high risks.

The STTGMA risk triage tool demonstrated an excellent level of accuracy in predicting fatal fall injuries. Earlier studies have reported the high discriminative ability of the STTGMA in predicting in-hospital mortality among older adults with hip fractures, and among patients who had orthopedic and neurosurgery consultations.(16, 17) Since hip fractures represent 10 to 15 percent of the complications that occur following fall injuries,(33) our study extended the utility of STTGMA for the larger geriatric fall-related trauma population.

Earlier studies have defined STTGMA risk categories either as three-level (minimal, moderate, and high-risk)(19, 20) or four-level (minimal, low, moderate, and high-risk)(21, 34) categorical variables. Our study showed that both approaches are effective depending on the outcome of interest. In assessing the hospital length of stay, the median number of days in the low and moderate-risk categories were not significantly different. The lack of discrimination between the low and moderate-risk categories may be due to the increased need for some older adults with fall injuries to be discharged to skilled nursing facilities as well as institutional policies that require a three-day criterion to be met before such discharge can be made. The three-day rule specifies that any patient that is to be discharged to a skilled nursing facility must have a medically necessary three consecutive days of in-hospital admission excluding the day of discharge and ED admission.(35)

While the exponential increase in mortality risk from minimal to low, moderate, and high-risk categories lends credence to the STTGMA triage tool’s predictive ability, it provides the opportunity to identify the at-risk population and create risk-specific interventions for such patient populations. Identifying community clusters of high fall mortality risk patient populations and implementing evidence-based interventions such as the Stopping Elderly Accidents, Deaths & Injuries (STEADI) (36-38) may reduce county-level and state-level fall-related mortality rates. Additionally, the Emergency Medical Service may embed the STTGMA risk scoring into their injury triage assessment and institute policies to ensure rapid transfer of older adults at risk of fatal fall injuries to levels I and II trauma centers.

This study has its limitations. Data entry errors, unreported history of falls, and inaccurately recorded history of background chronic medical conditions may influence the results of this study. Additionally, our study is from a single institutional trauma database and our result may not be generalizable to other trauma centers. The STTGMA risk triage scoring is computationally complex, and this may deter physicians and other providers from using this powerful predictive tool. Healthcare institutions may subvert this challenge by integrating the publicly available tool into their electronic health record system and automating the risk calculations for trauma patients 55 years and older. Since the risk categorization is defined using percentiles, every healthcare system that wishes to integrate the STTGMA into its departmental workflow will have to use its retrospective data as a primer to generate the STTGMA risk categories. However, the ability to adapt the STTGMA risk scoring to different patient populations is one of the unique qualities of the risk triage tool. This study addresses the need for a highly predictive risk triage tool for geriatric trauma patients and it is the first to report the predictive accuracy of the STTGMA risk triage tool among older adults with fall injuries.

## Conclusion

STTGMA predicts in-hospital mortality from fall injuries with high accuracy and can risk-stratify and predict hospital length of stay and time to death from fall injuries in older adults. The ability to identify older adults likely to have fatal outcomes from fall injuries can guide in-hospital care protocols, and design interventions to target this at-risk population.

## Data Availability

All data produced in the present study are available upon reasonable request to the authors

## Data Availability

The data that support the findings of this study are available on request from the corresponding author.

